# Scarce COVID-19 Testing Capabilities at Urgent Care Centers in States with Greatest Disease Burden

**DOI:** 10.1101/2020.03.22.20040923

**Authors:** Walter Hsiang, Howard Forman, Siddharth Jain, Akshay Khunte, Grace Jin, Laurie Yousman, Michael Najem, Alison Mosier-Mills, Daniel Wiznia

**Affiliations:** Yale School of Medicine, New Haven, CT; Yale School of Medicine, Department of Radiology, New Haven, CT; Yale School of Medicine, Department of Orthopedics and Rehabilitation, New Haven, CT

## Abstract

As of March 22, 2020, the number of confirmed COVID-19 cases in the U.S. has reached nearly 30,000.^1^ While rapid and accessible diagnosis is paramount to monitoring and reducing the spread of disease, COVID-19 testing capabilities across the U.S. remain constrained. For many individuals, urgent care centers (UCCs) may offer the most accessible avenue to be tested. Using a phone survey, we describe the COVID-19 testing capabilities of UCCs in states with the greatest disease burden.

## Methods

We identified ten states with the highest COVID-19 caseload as of March 19 according to the Centers for Disease Control (CDC).^2^ 25 UCCs, defined as walk-in clinics in ambulatory medical facilities, were randomly selected and called from each state using the Urgent Care Association “Find an Urgent Care” directory. UCCs were classified into independent, hospital/health network, and academic categories.

Using a standardized survey script (Figure 1), trained investigators asked UCC receptionists about COVID-19 testing ability, testing criteria, time to test results, costs of tests and visits for insured/uninsured patients, and test referrals. All 250 calls were made on March 20 and were limited to 1 minute to minimize occupying clinic resources.

**Figure 1:**
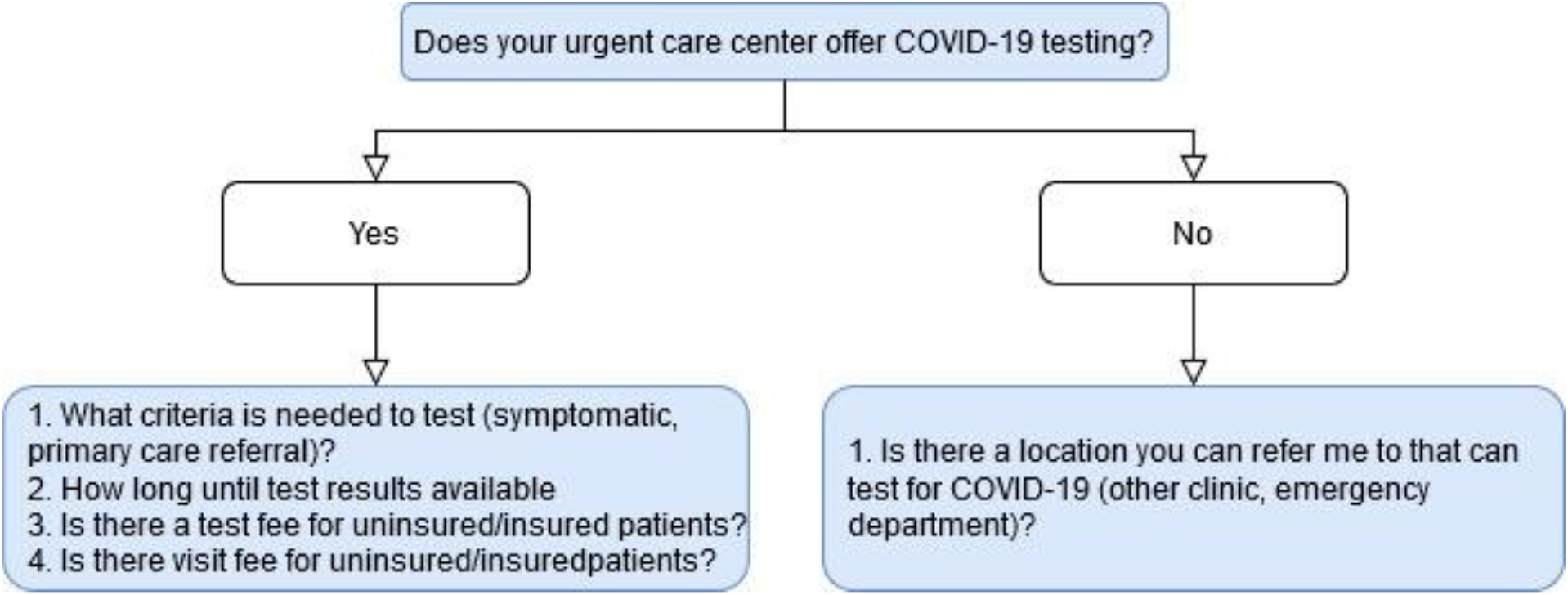
Urgent care center survey script.

## Results

Of 250 UCCs contacted, 57 (22.8%) offered COVID-19 testing. Hospital/health network-affiliated UCCs were more likely to offer COVID-19 tests compared to independent UCCs (odds ratio 3.69, 95% confidence interval 1.94-7.01, p<0.0001).

Of UCCs that offered testing, 56 (98.2%) required the patient to be symptomatic (typically fever and respiratory symptoms) and 2 (0.4%) required a primary care physician referral. 45 (86.5%) UCCs charged a fee to test uninsured patients, but no UCC could provide a definitive answer regarding test fees for insured patients given the shifting federal legislation. 53 (94.6%) UCCs charged a visit fee in addition to the COVID-19 lab test fee. For the 49 centers that provided the wait time for test results, the median time was 120 hours (interquartile range 96 hours to 144 hours).

Of UCCs that did not offer testing, 97 (51.3%) referred individuals to other clinics that could possibly test for COVID-19, and 37 (24.8%) directly referred individuals to a specific emergency department.

## Discussion

In the ten states with the greatest COVID-19 caseload, only 23% of UCCs offered COVID-19 testing. Additionally, results would take approximately five days to be processed. Although time to test results at public/state labs are typically 24-48 hours (Table 1), time to test results at UCCs were longer as most samples are sent to external labs. However, it remains unclear whether UCC ability to obtain test samples may be unmatched by the ability to process tests. This finding underscores the importance of point-of-care testing that can rapidly detect COVID-19, particularly because severe disease peaks at approximately ten days from onset of initial symptoms.^3^

**Table 1:** COVID-19 testing capabilities by state.

|  | UCCs offering tests, n (%) | Average time to test results at UCC, hours | Average time to test results at state or public health lab, hours* |
| --- | --- | --- | --- |
| **California** | 1 (4%) | N/A | 48-72 |
| **Colorado** | 2 (8%) | 108 | 72 |
| **Florida** | 7 (28%) | 96 | 24-48 |
| **Georgia** | 2 (8%) | 120 | N/A |
| **Illinois** | 6 (24%) | 118 | 24 |
| **Louisiana** | 4 (16%) | 138 | N/A |
| **Massachusetts** | 9 (36%) | 139.2 | 24 |
| **New Jersey** | 7 (28%) | 124.5 | 24-48 |
| **New York** | 10 (40%) | 115.2 | 3-5 |
| **Washington** | 9 (36%) | 91 | 24-48 |

\*Time to results at state/public health labs obtained from the respective state's Department of Public Health website as of March 20.

Fees and cost-sharing for COVID-19 tests remain unclear. The Families First Coronavirus Response Act, which passed on March 18, mandated all group and individual health plans cover COVID-19 testing and gave states the option to use Medicaid coverage for testing uninsured patients.^4^ Although this study could not definitively define test fees, most UCCs stated they would charge test fees, contrary to recent federal regulations, in addition to fees for the urgent care visit itself as of March 20. Test and visit fees at UCCs may discourage patients from seeking COVID-19 testing.

This report has limitations. The small number of UCCs contacted per state may not accurately represent the state’s urgent care climate. Additionally, the rapidly changing nature of the COVID-19 pandemic may affect these findings. However, this study serves as an important snapshot that highlights the limited COVID-19 testing capabilities at UCCs in the most heavily burdened states.

## Data Availability

Data is available to view or use via request to the principal investigator.

## Abbreviations

UCC: (urgent care center)

